# Second-Hand Smoke Exposure inside the House and Adverse Birth Outcomes in Indonesia: Evidence from Demographic and Health Survey 2017

**DOI:** 10.1101/2021.11.20.21266641

**Authors:** Helen Andriani, Nurul Dina Rahmawati, Abdillah Ahsan, Dian Kusuma

**Author notes:** Corresponding author: Helen Andriani, Ph.D. Address: Kampus Baru UI Depok, Depok 16424, Indonesia, Department of Health Policy and Administration, Faculty of Public Health, Universitas Indonesia, Depok, Indonesia.

## Abstract

**Objectives:** Second-hand Smoke (SHS) during pregnancy among non-smoking women associates with mortality and morbidity risks in their infants. However, little is known about the SHS inside the house and the adverse birth outcomes. This study aims to assess the prevalence, level, and frequency of SHS exposure inside the house and investigate their associations with birth outcomes.

**Methods:** We use the Indonesian Demographic and Health Survey (IDHS) 2017, a large-scale nationally representative survey. Women aged 15 to 49 years who had given birth in the last five years before the study and their husbands were interviewed (n=19,935). Three dependent variables included Low Birth Weight (LBW), size at birth, and birth weight.

**Results:** Seventy-eight percent of mothers exposed to SHS inside home, of whom 7.2% had LBW children. Compared to non-SHS exposure mothers, those exposed to SHS were younger, had first birth before 20 years old, married, lower educated, non-worker, lived in rural, grand multipara, had pollution from cooking fuel, cook in a separate building, had higher risk of delivering lower birth weight (aOR=1.16, 95% CI: 1.02, 1.33), and smaller baby (aOR=1.51, 95%CI: 1.35, 1.69), even after the controlling for the covariates. We identified the inverted U-shaped association for SHS exposure frequency. Similar risk was also observed among mothers exposed with SHS on a daily basis compared to those who are not exposed.

**Conclusion for Practice:** Exposure to SHS inside home was significantly associated with LBW and size at birth. Given the high smoking prevalence, relevant policy and health promotion are needed.

**Significance Statement:** Adverse birth outcomes, such as low birth weight and smaller size at birth, may not be clearly explained by the second-hand smoke (SHS) exposure from a smoking husband alone. Our findings show that the prevalence of SHS inside the house in Indonesia is 78.4%. Pregnant women exposed to anyone who smokes in the household may be linked to poor birth quality, including low birth weight and smaller size at birth in their babies, after adjustment for risk factors. The effects of SHS exposure on birth outcomes are further exacerbated by daily SHS exposure.

## Introduction

Globally 600,000 deaths are attributable to Second-hand Smoke (SHS) in each year (Oberg et al., 2011) and more than 33% of the population is often actively or passively exposed to cigarette smoke and about 35% of all female non-smokers are exposed to SHS (Data). Tobacco smoke exposure and the resulting inhalation of SHS is among the most widespread forms of micro-environmental exposure (i.e., indoor) is among the most widespread forms of micro-environmental exposure (i.e., indoor). Women and children under five years of age are most vulnerable in the houses or indoor air pollution. A retrospective study from 192 countries reported that 40% of children were exposed to SHS, higher than non-smoking men and women, that were only 33% and 35%, respectively, while 47% of death contributed to SHS occurred in women, almost twice higher than in children and men (Oberg et al., 2011). In addition, 36% of children even got exposed since they were in the womb (Cheng et al., 2017). At least 40% of children had become passive smokers because of SHS in their homes, and 31% of them die due to the cigarette smoke they breathe every day (Oberg et al., 2011).

SHS during pregnancy among non-smoking women is associated with mortality and morbidity risks in their infants, including stillbirth, prematurity, miscarriage, and Low Birth Weight (LBW) (Jaddoe et al., 2008). SHS also increased risk of stunting (length for age), wasting (weight for age), and underweight (weight for length) among children as well as smaller head circumference resulting smaller babies, on the other hand increased susceptibility of being overweight at the same time, in accordance to the Gross National Income of the country (Nadhiroh et al., 2020). Evidence in Indonesia showed that compared to non-exposed pregnant women, the risk of having low birth weight infants were around three times higher in exposed ones. Similar risk was also drawn based on the number of active smokers at home, number of cigarettes consumed, and exposure duration in a daily basis (Ardelia et al., 2019).

Based on Demographic and Health Survey data collected between 2008 and 2013 from 30 Low-income and Middle-income Countries (LMICs), daily SHS exposure accounted for a greater population attributable fraction of stillbirths than active smoking was 14% in Indonesia. This number is the highest among other 30 LMICs (S. Reece et al., 2019). Indonesia has compiled various regulations governing public protection from the dangers of exposure to cigarette smoke. One of them is through the adoption of no-smoking zones in various public places and workplaces, especially in schools and hospitals. However, World Health Organization (WHO) notes that regulations regarding smoke-free areas in public areas in Indonesia are still relatively low compared to other Southeast Asia countries and in accordance with the geographic distribution as well as socio-economic disparity, where urban settings, wealthier and more educated population were more likely adopt smoke-free policy (Wahidin et al., 2020).

Given the implications for child mortality, a significant reduction in prevalence of LBW is necessary in order to achieve Sustainable Development Goals (SDGs) and a similar need to strengthen the implementation of the Framework Convention on Tobacco Control (FCTC) of the WHO in all countries (Organization, 2015a). Only few robust studies examined clear association between the exposure to SHS inside the house and birth outcomes, especially in Indonesia (Noriani et al., 2015; Sian Reece et al., 2019; Soesanti et al., 2019). This study contributes to fill knowledge gap in SHS exposure inside the house and adverse birth outcomes in Indonesia, using the latest and quantitative evidence of a large-scale population-based data and taking into account SHS frequency, LBW, and size at birth, neither of which have been presented in previous studies. This study aims to assess the prevalence, level, and frequency of SHS exposure inside the house and investigate the association between SHS exposure inside the house and birth outcomes.

## Methods

### Data sources

We use data from the latest survey of Indonesia Demographic and Health Survey (IDHS) 2017, a nationally representative, large-scale, and repeated cross-sectional household survey collecting population, health, and nutrition data. All ever married women between 15 to 49 years of age who had given birth in the last five years prior to the survey in sampled households are eligible for an interview using a standard self-reported questionnaire (Population et al., 2018). The reason women were chosen to give birth during the last five years prior to the survey was to prevent bias in memory recall from mother. A written, informed consent was given to all participants. The total sample size in the study was 19,935.

### Ethical oversight

The 2017 IDHS program collects data periodically for policy development and program planning, monitoring and evaluation. Respondents read an informed consent statement before each interview. The statements include voluntary participation, refusal to answer questions or termination of participation at any time and confidentiality of identity and information. The Institutional Review Board (IRB) of the Inner City Fund International Inc., Fairfax, VA, USA reviewed and approved the study procedures and survey protocols. After obtaining authorization from the IDHS to use the dataset, the IRB of Universitas Indonesia provided further ethical review approval (598/UN2.F10.D11/PPM.00.02/2020).

### Measurement

Two main independent variables were the exposure to SHS inside the house and SHS exposure frequency. A binary variable (not exposed vs exposed) was created to measure exposure to SHS inside the house, where one or more adult smoke commercial cigarettes, cigars, and other country-specific smoking products. SHS smoking frequency was classified as: not exposed, less than once a month, monthly, weekly, and daily.

Three outcomes variable related to the self-reported birth outcomes are: 1. LBW, 2. Size at birth, and 3. Birth weight. We treated LBW (<2,500; >2500 grams) and size at birth (mothers’ perception of their newborns’ size at birth: smaller than average; average; and larger than average) as categorical variables. We note that there are mixed results in the literature on the validity of using mother’s perceptions on size (Channon, 2011; Nigatu et al., 2019), but for our data we found significant positive association of the perceived size and birth weight (see Table S1). Birth weight (in grams) was treated as continuous variable. Control variables included demographic and socioeconomic characteristics such as maternal age, age at first birth, marital status, maternal education, family size, mother’s occupation, husband’s education, residence (urban; rural), parity, birth interval, birth order, wealth index (a composite measure of a household’s cumulative living standard or ownership of selected assets), cooking fuel, and kitchen location.

### Statistical analyses

Data were analyzed using SPSS version 25. The proportions and chi-squared tests were used to test for differences between SHS exposure inside the house, demographic and socioeconomic characteristics. Logistic regression analyses were conducted to measure relative odds of associations of SHS exposure and frequency inside the house with LBW and size at birth. Using General Linear Model, we assessed the relationships of SHS exposure and frequency inside the house with birth weight. All multivariable models were used to control for background characteristics. In addition to significant covariates, backward elimination was performed as the variable selection procedure to retain important confounding variables, resulting potentially in a slightly richer model. The overall model is also evaluated using the goodness of fit test by likelihood ratio test.

## Results

The characteristics of the participants are presented in Table 1. Seventy-eight percent of the mothers in our study were exposed to SHS in the household, with 7.2% of those who had LBW being exposed SHS. Compared with non-SHS exposure mothers, mothers exposed to SHS were aged 15-24 years, had first birth before 20 years of age, married, had a lower education, non-worker, lived in a rural area, grand multipara, had pollution from cooking fuel, and cook in a separate building. All the indicators were statistically significant at p<0.05, except for husband’s occupation and birth interval, which were not different in exposure to SHS.

**Table 1.**
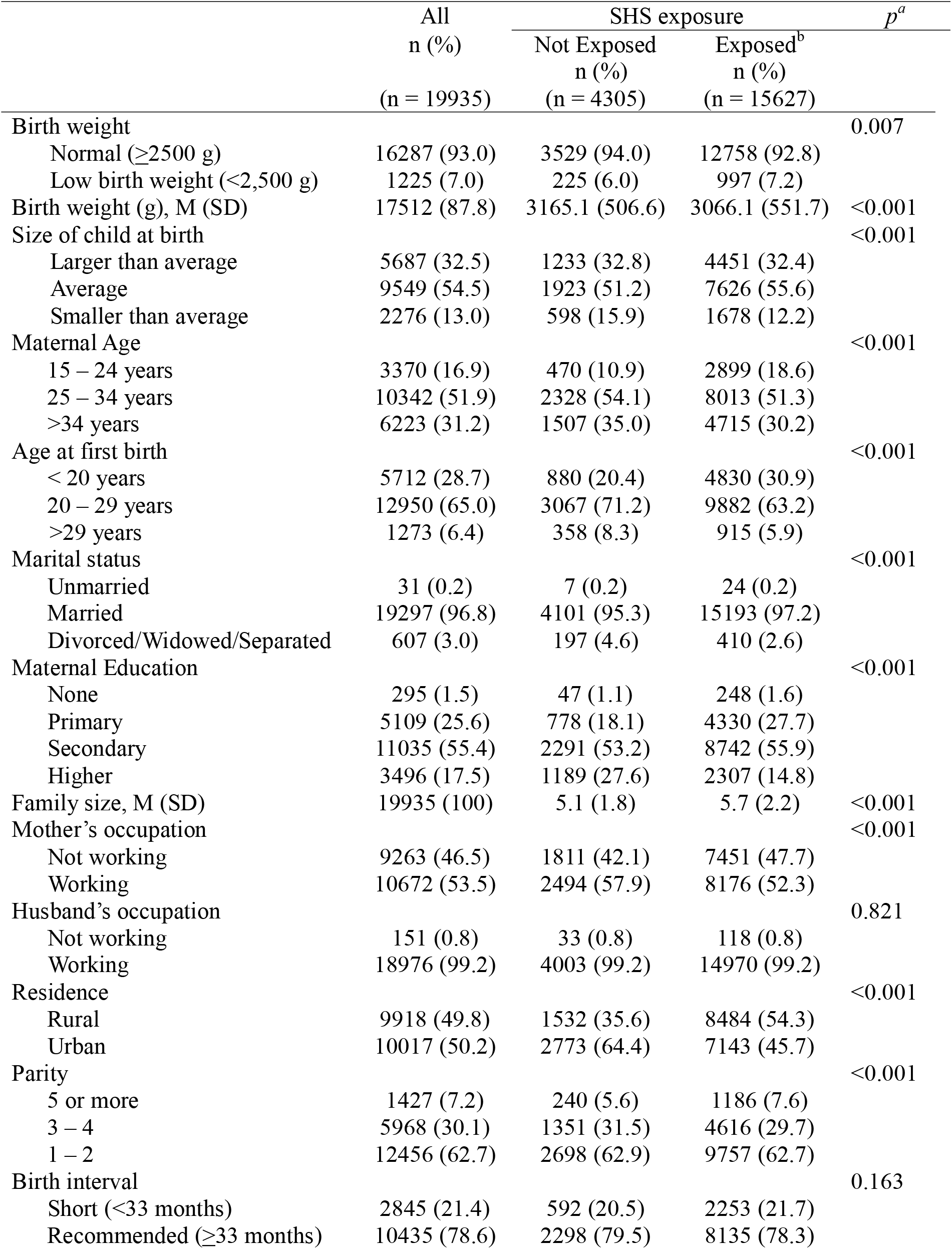
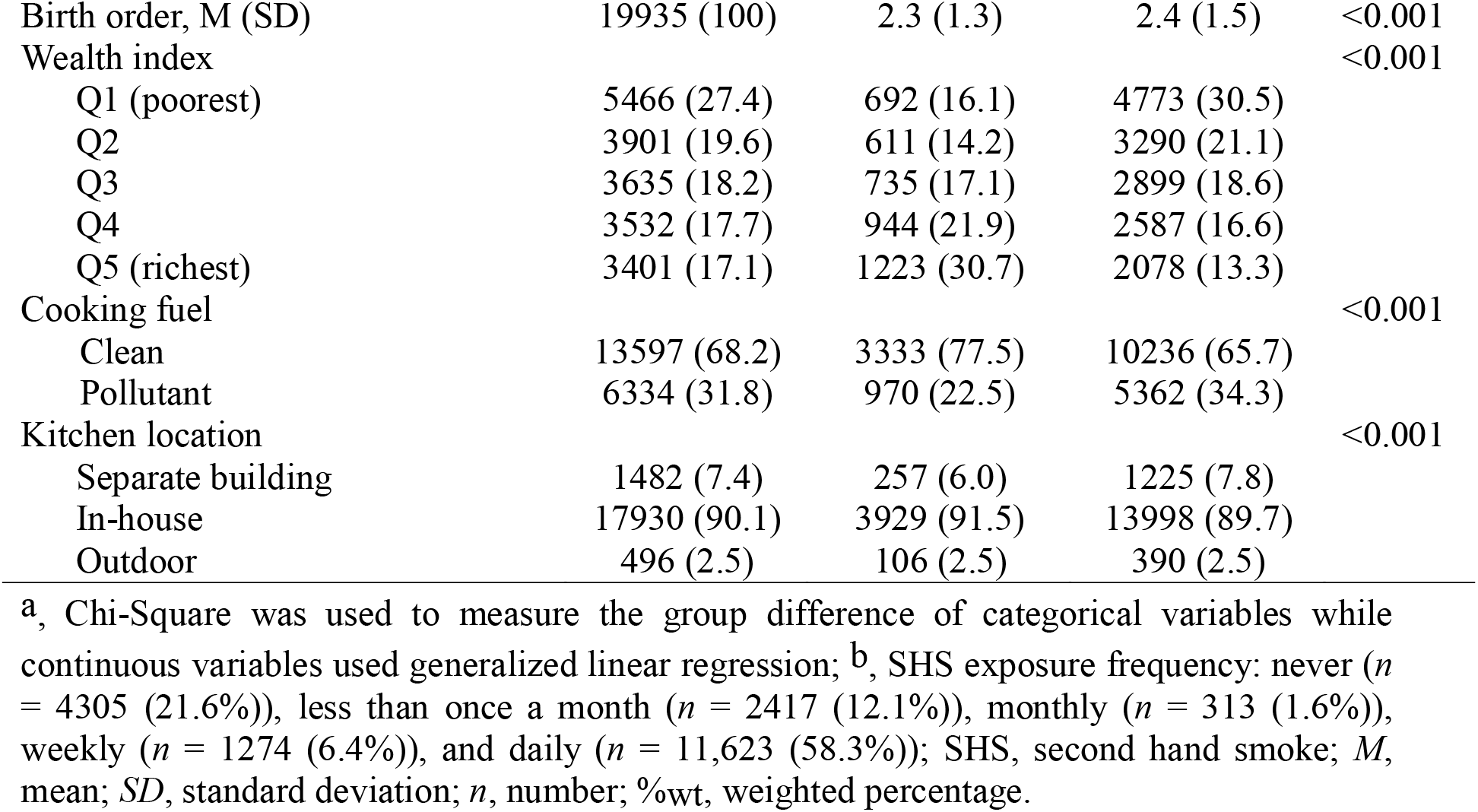
Characteristics of participants by exposure to SHS inside the house

Table 2 shows the association between SHS exposure inside house and birth weight and LBW. The mean birth weight was significantly associated with SHS exposure inside the house. After adjusting for the covariates, mothers exposed to SHS had children with a mean birth weight of 71,6 g (p<0.001) lower than that of mothers who did not exposed to SHS. SHS exposure significantly raised the odds of having LBW children (OR=1.21, p<0.05). After adjusting for the covariates, compared to non-SHS exposure, mothers who exposed to SHS showed a significant 1.16-fold increase in the odds for having LBW children (adjusted OR=1.16, 95% CI [1.02, 1.33], p<0.050). For SHS exposure frequency, mothers exposed to SHS daily had children with a mean birth weight of 63,4 g (p<0.001) lower than that of mothers who did not exposed to SHS. SHS exposure weekly and daily significantly raised the odds of having LBW children (OR=1.37, p<0.050 and OR=1.21, p<0.050, respectively). After adjusting for the covariates, compared to non-SHS exposure, mothers who exposed to SHS weekly and daily showed a significant 1.33-fold and 1.18-fold increase in the odds for having LBW children (adjusted OR=1.33, 95% CI [1.03, 1.71], p<0.050 and adjusted OR=1.18, 95% CI [1.01, 1.38], p<0.050). We have identified the inverted U-shaped association between SHS exposure frequency and birth weight or LBW. Daily exposure group showed a statistically significant lower adjusted mean of birth weight than non-exposed group and lower odds of LBW than weekly exposure group, but still showed a higher adjusted mean of birth weight when compared with weekly or other categories of SHS and a higher odd of LBW when compared with non-exposed or other categories of SHS.

**Table 2.**
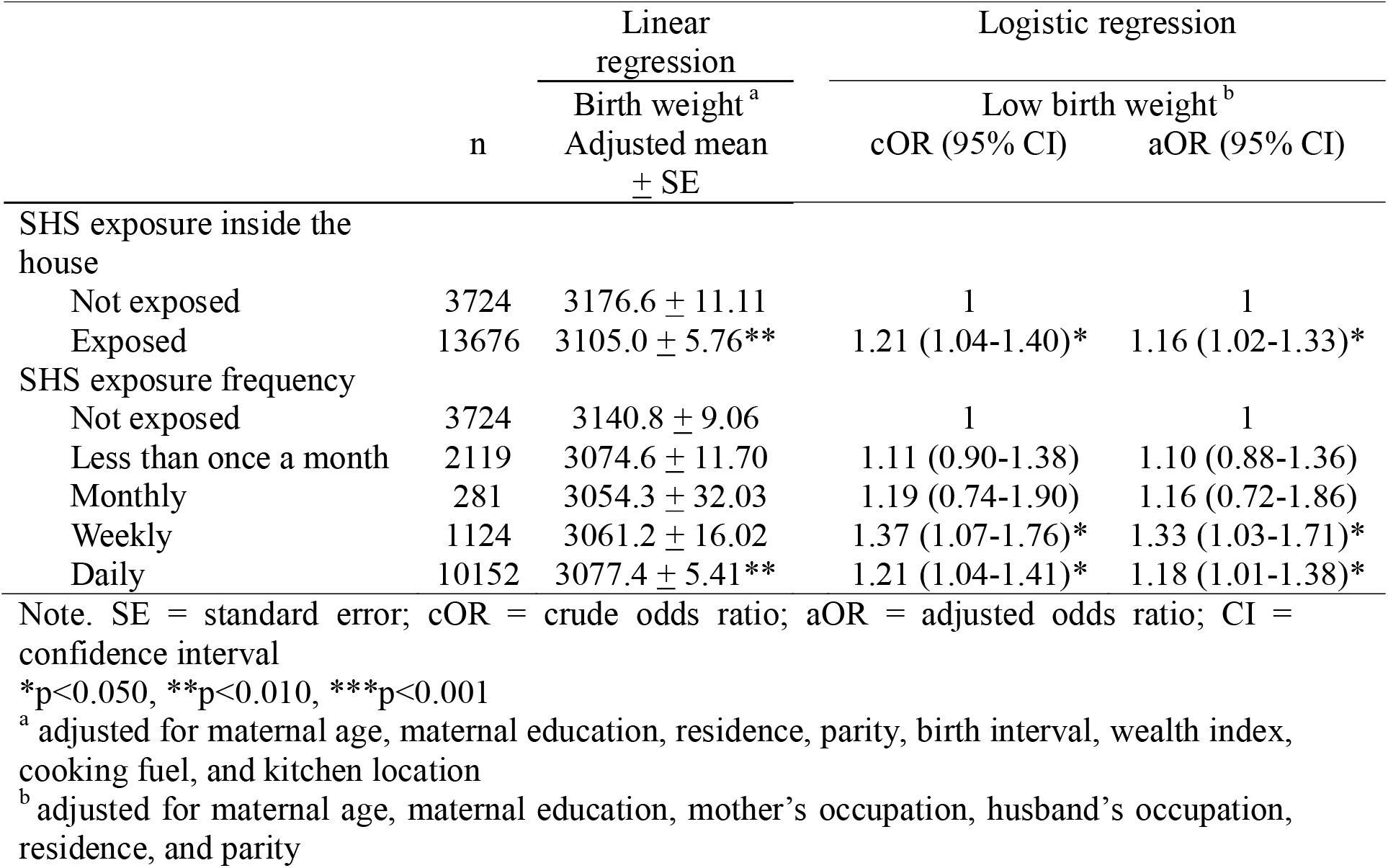
Association between SHS exposure inside the house and birth weight and low birth weight

Table 3 shows the association between SHS exposure size at birth, using multinomial logistic model. Mothers who exposed to SHS significantly raised the odds of having larger than average size at birth of their children (OR: 1.09, p<0.05) and having smaller than average size at birth of their children (OR: 1.41, p<0.001). After adjusting for the covariates, compared to those who did not expose to SHS, mothers who exposed to SHS showed a significant 1.10-fold increase in the odds for having larger than average size at birth of their children (adjusted OR (95% CI): 1.10 (1.01 – 1.20), p<0.05) and having smaller than average size at birth of their children (adjusted OR (95% CI): 1.51 (1.35 – 1.69), p<0.001). Mothers who exposed to SHS daily significantly raised the odds of having larger than average size at birth of their children (OR: 1.10, p<0.05) and having smaller than average size at birth of their children (OR: 1.43, p<0.001). After adjusting for the covariates, compared to those who did not expose to SHS, mothers who exposed to SHS daily showed a significant 1.11-fold increase in the odds for having larger than average size at birth of their children (adjusted OR (95% CI): 1.11 (1.02 – 1.201), p<0.05) and having smaller than average size at birth of their children (adjusted OR (95% CI): 1.54 (1.37 – 1.73), p<0.001).

**Table 3.** Association between SHS exposure inside the house and size at birth (multinomial logistic regression)

|  | n | Size at Birth |  |  |  |
| --- | --- | --- | --- | --- | --- |
|  |  | Larger than average†<sup>a</sup> |  | Smaller than average†<sup>a</sup> |  |
|  |  | cOR | aOR (95% CI) | cOR | aOR (95% CI) |
| SHS exposure inside the house<sup>a</sup> |  |  |  |  |  |
| Not exposed | 3708 | 1 | 1 | 1 | 1 |
| Exposed | 13656 | 1.09* | 1.10 (1.01-1.20)* | 1.41*** | 1.51 (1.35-1.69)*** |
| SHS exposure frequency<sup>b</sup> |  |  |  |  |  |
| Not exposed | 3708 | 1 | 1 | 1 | 1 |
| Less than once a month | 2122 | 0.97 | 0.97 (0.84-1.11) | 1.07 | 1.08 (0.90-1.31) |
| Monthly | 281 | 0.89 | 0.88 (0.67-1.16) | 1.19 | 1.25 (0.88-1.77) |
| Weekly | 1116 | 1.06 | 1.07 (0.96-1.18) | 1.00 | 1.04 (0.89-1.21) |
| Daily | 10137 | 1.10* | 1.11 (1.02-1.21)* | 1.43*** | 1.54 (1.37-1.73)*** |

Note: cOR = Crude Odds Ratio; aOR = Adjusted Odds Ratio; CI = Confidence Interval
†Average is the reference category
\*p<0.05, \*\*p<0.01, \*\*\*p<0.001
<sup>a</sup>adjusted for maternal age, age at first birth, maternal education, family size, mother's occupation, residence, parity, birth order, wealth index, cooking fuel

## Discussion

Our findings show that 78.4% of study sample were exposed to SHS inside the house, which is remarkably higher than the prevalence of SHS exposure at home from other countries such as China (48.3%), Bangladesh (46.7%), and Thailand (46.8%) (Fischer et al., 2015; Phetphum & Noosorn, 2020; Wang et al., 2009). SHS is a major source of indoor pollution of particulate matter and is further enhances inside the houses by cigarette smoking. Cigarette smoke contains toxic substances and has detrimental effects on almost every organ in the body. Besides, tobacco smoke contains a large variety of poisonous gasses and particles that are hazardous to not only the smokers but also to those around them (Nafees et al., 2012).

We also found significant associations between SHS exposure and birth weight, LBW, and size at birth. SHS exposure in the house during pregnancy raised the risk of LBW, which was 7.2% more likely in household for babies whose mothers were exposed to SHS compared to babies whose mothers were not. The frequency of exposure has also contributed significantly to LBW risk. As the prevalence of smoking is rising in some Asian countries due to a change in tobacco markets from high to low-income countries (Organization, 2015b), adverse birth outcomes such as LBW and size at birth may be unavoidable. Our research showed that mothers exposed to SHS in Indonesia were 1.16 times more likely to have LBW newborns and 1.51 times more likely to have smaller infants. In terms of SHS exposure frequency, mothers who were exposed to SHS at a daily rate were at an increased risk of having LBW newborns and smaller infants. The results of other studies in different populations, such as the Netherlands, USA, and Greece have also shown that exposure to SHS is positively correlated with LBW risk (Jaddoe et al., 2008; La Merrill et al., 2011; Vardavas et al., 2010).

Previous retrospective and prospective cohort studies confirmed that the expose of domestic cigarette smoke throughout the pregnancy was significantly related to the lower adjusted mean birth weight as well as doubling the risk of having smaller baby (Kobayashi et al., 2019; Norsa’adah & Salinah, 2014). Voigt et al. also reported that the mean birth weight of exposed group was significantly lower no matter the Body Mass Index (BMI) of the mothers is (Voigt, Jorch, et al., 2011). It was suggested that there was 12.9 g reduction of birth weight related to exposure of each additional cigarette smoke and prevalence of LBW was twice higher in exposed compared to non-exposed women (Norsa’adah & Salinah, 2014). The plausible mechanism for the increased risk of LBW can be explained by two pathways, namely: 1) the elevation of Tumor Necrosis Factor alpha triggered by SHS exposure which is transmitted across placenta and might directly increase the risk of LBW, and 2) significant reduction of placental weight by almost 4 times lower compared to un-exposed mothers due to the increased secretion of TNF-α and Vascular Cell Adhesion Molecule-1 as inflammatory markers. These factors can be directly associated with LBW since the abundant level of inflammatory markers in pregnant women lead to the damage of placenta and thus nutrient and oxygen cannot be optimally delivered to the fetus, resulting suboptimal growth and LBW (Niu et al., 2016).

The inverted U-shaped association were likely attributable to the age of mother, education, occupation, parity, and wealth index. Compared with non-daily exposure, mothers exposed to daily SHS were aged 15-24 years, had first birth before 20 years of age, had a lower education, non-worker, grand multipara, and were from lower wealth index. There were 58% of women in our sample had daily exposure; these women were younger maternal age (the proportion of women maternal age 15-24 was 12.7% vs 19.9% with non-daily vs daily (or 1.6 times)), which may drive the higher birth rates among daily. One study in China showed that birth weight increased 16.204 g per year when maternal age was less than 24 years old, and increased 12.051 g per year when maternal age ranged from 24 to 34 years old, then decreased 0.824 g per year from 34+ years (Wang et al., 2020). In our study, although it has been controlled for maternal age in regression, it still shows U-shaped for daily, so that the potential explanation could be maternal age. Smoking prevalence in men will raise the risk of SHS exposure among young women who have never smoked (Lim et al., 2018). Since many women spend the majority of their time at home, the home is the most common source of SHS exposure for unemployed women. Well-educated and higher-income women may have a greater understanding of the hazards of SHS exposure and may have more positive attitudes toward SHS exposure in the home, resulting in them becoming more conscious of avoiding SHS (Sun et al., 2016). Also, because younger maternal age may tend to be lower education, employability, income, and higher parity. The self-administration of nicotine (positive re-enforcing effect) need optimum doses near the top of the inverted U that may enhance growth, longevity, etc., but at higher doses, aversive effects dominated (Heishman et al., 2010). These findings reinforced the social inequality associated with smoking, backed up the need for further research on the subject, and emphasized the need for more approaches to control the issue.

In terms of birth size, the exposure of SHS attenuated the risk of delivering macrosomia baby (weighted ≥4,000 g) by almost 2 times among obese women (Body Mass Index of ≥30 kg/m^2^) and on the other hand increasing the risk of delivering LBW infants among underweight women (BMI of <18 kg/m^2^) to almost 3-fold (Wahabi et al., 2013). The nicotine exposure during pregnancy impaired the differentiation and proliferation of placenta cell named cytotrophoblast (CTB) that reduce the blood flow and create pathological hypoxic environment in the womb (Zdravkovic et al., 2005). Voigt et al. in another study suggested that the prevalence of Small Gestational Age (SGA) defined by birth weight, length, and head circumference is elevating in accordance with the increasing of cigarette smoke exposure up to 8% for each seven additional cigarettes daily (Voigt, Zels, et al., 2011).

Policymakers should align tobacco control initiatives with maternal, newborn, and child health care services with continued evidence of the health effects of the SHS. Indonesia’s cultural norms have also attributed to exposure to SHS in houses, as older people are not required to avoid smoking in homes. The limited awareness and knowledge about the health risks of SHS among active and passive smokers is also a key factor. Passive smoking seems to be a neglected public health problem in Indonesia. Cigarette smokers should be forced to stop indoor smoking, and smoking cessation campaign services should also be strengthened. To reduce exposure to SHS, enforcing strict tobacco control laws and promoting smoke-free environments is also crucial, as is encouraging mothers to take precautionary action to avoid exposure to SHS during pregnancy. The attitude of parents and health-related behaviors that control SHS exposure to non-smokers, can be influenced by electronic and social media. Moreover, it can be used to raise awareness and provide information on the hazards of SHS and passive smoking. Preventing family members from smoking inside the houses and creating smoke-free homes can also act as a powerful strategy of tobacco control for adolescents (Song et al., 2009).

There were many limitations in our study. A self-reported questionnaire of SHS exposure and frequency without measurement of the duration of maternal or pregnancy exposure and cotinine levels in the household may have been underestimated the true impact of SHS in Indonesia. Also, we are unable to match the period of smoking experience (by anyone) and the period of pregnancy. Secondly, since we used a cross-sectional dataset and worked under the presumption that SHS was even over time, our results should be interpreted with caution, and thus causality could not be determined. With reliable measurement of exposure to SHS toxins, a prospective cohort study would be suitable. Self-reported variables may lead to misclassifications due to recall and reporting bias (i.e., size at birth reported by the mother may prone to subjectivity). Only crude categories (exposed vs not exposed) were selected for the multivariable analysis to prevent any false exposure categories precision. Finally, we were unable to account for a variety of unmeasured confounders, such as biological influences, including food intake and nutritional status of pregnant women, which may clarify our results because for general national policy purposes, the data was collected and is therefore limited. Nonetheless, our research contributes to the body of knowledge on the birth effects of SHS with regards to birth weight and size at birth. With a wide range of representative populations in Indonesia, this is the first study of its kind, thereby raising the generalizability of our results to the Indonesian population, the largest cigarette consumer in the Asia Pacific region.

In conclusion, household exposure to SHS remains a relevant risk factor and is significantly associated with lower birth weight, LBW, and smaller size at birth at population level in Indonesia. For the implementation of strategies to minimize SHS exposure, the home of non-smokers living with smokers must be taken into account through smoke-free homes policy (Trisnowati et al., 2019). Therefore, it is not only important to enact regulations, but also to consider more public health strategies to raise awareness of the adverse health effects caused by SHS exposure. It is important to do further research to investigate the duration of SHS exposure and other biological plausibility. Public health promotions that disseminate general recommendations for action are likely to have a positive impact on the health of mothers, newborns, and children.

## Data Availability

Data are available upon reasonable request. The data are available from the Indonesia Demographic and Health Survey (IDHS), collected in 2017. The dataset is publicly available at IDHS website (https://dhsprogram.com/data/dataset/Indonesia_Standard-DHS_2017.cfm?flag=0). The author granted access to the dataset by registering for access to the IDHS data download link.

## Declaration

### Ethics approval

The Institutional Review Board (IRB) of the Inner City Fund International Inc., Fairfax, VA, USA reviewed and approved the study procedures and survey protocols. After obtaining authorization from the IDHS to use the dataset, the IRB of Universitas Indonesia provided further ethical review approval (598/UN2.F10.D11/PPM.00.02/2020).

### Consent to participate

Not applicable

### Consent for publication

Not applicable

### Availability of data and material

Not applicable

### Code availability

Not applicable

### Authors’ contributions

Conceptualization: HA and NDR; Data acquisition and curation: HA; Methodology: HA, NDR, DK; Software: HA; Formal analysis: HA; Interpretation: HA, NDR, AA, DK; Writing original draft: HA and NDR; Writing critical review: HA, NDR, AA, DK. All authors read and approved the final manuscript.

## Acknowledgments

The authors would like to acknowledge the Indonesian Demographic and Health Surveys (IDHS) Program for the access to the data used in this study. This study was supported by the research grant from Indonesian Tobacco Control Research Network (ITCRN) 2020, in collaboration with the Center of Islamic Economics and Business, Faculty of Economics and Business, Universitas Indonesia and the Johns Hopkins Bloomberg School of Public Health. The funders had no role in the design of the study and collection, analysis, and interpretation of data and in writing the manuscript.

