## Supplementary material for "Second-Hand Smoke Exposure inside the House and Adverse Birth Outcomes in Indonesia: Evidence from Demographic and Health Survey 2017": Table S1

**Table S1.** Association between birth weight and size at birth

The mothers' perceptions of birth size are positively correlated with birth weight (in grams) and birth weight status (normal vs LBW)

|  | Size at Birth | | | | *p* |
| --- | --- | --- | --- | --- | --- |
|  | Larger than average  n (%)  (n = 5687) | Average  n (%)  (n = 9549) | Smaller than average  n (%)  (n = 2276) |  | |
| Birth weight  Normal  Low birth weight | 5687 (100.0)  0 (0.0) | 8799 (92.1)  750 (7.9) | 1801 (79.1)  475 (20.9) | <0.001 | |
| Birth weight (g), M (SD) | 3522.6 (6.2) | 2959.6 (4.1) | 2535.0 (9.8) | <0.001 | |

**Table S2.** Characteristics of participants by daily vs non-daily SHS exposure inside the house

|  | SHS exposure | | *p* |
| --- | --- | --- | --- |
|  | Non-daily ^a^  n (%)  (n = 8309) | Daily  n (%)  (n = 11623) |  |
| Maternal Age  15 – 19 years  20 – 24 years  25 – 29 years  30 – 34 years  >34 years | 123 (1.5)  934 (11.2)  2115 (25.5)  2355 (28.3)  2782 (33.5) | 305 (2.6)  2007 (17.3)  2898 (24.9)  2973 (25.6)  3440 (29.6) | <0.001 |
| Age at first birth  < 20 years  20 – 29 years  >29 years | 1930 (23.2)  5737 (69.0)  642 (7.7) | 3780 (32.5)  7212 (62.0)  631 (5.4) | <0.001 |
| Maternal Education  None  Primary  Secondary  Higher | 120 (1.4)  1739 (20.9)  4444 (53.5)  2006 (24.1) | 175 (1.5)  3369 (29.0)  6589 (56.7)  1490 (12.8) | <0.001 |
| Mother’s occupation  Not working  Working | 3600 (43.3)  4709 (56.7) | 5662 (48.7)  5961 (51.3) | <0.001 |
| Parity  5 or more  3 – 4  1 – 2 | 500 (6.0)  2606 (31.5)  5169 (62.5) | 926 (8.0)  3361 (29.0)  7286 (63.0) | <0.001 |
| Wealth index  Q1 (poorest)  Q2  Q3  Q4  Q5 (richest) | 1714 (20.6)  1391 (16.7)  1442 (17.4)  1683 (20.3)  2079 (25.0) | 3751 (32.3)  2510 (21.6)  2192 (18.9)  1848 (15.9)  1322 (11.4) | <0.001 |

^a^ Non-daily: never, less than once a month, monthly, and weekly

The prevalence of daily exposure to SHS at home was 58.3%. Compared with non-daily exposure, mothers exposed to daily SHS were aged 15-24 years, had first birth before 20 years of age, had a lower education, non-worker, grand multipara, and were from lower wealth index.
